# Ambient heat exposure and COPD hospitalisations in England: A nationwide case-crossover study during 2007-2018

**DOI:** 10.1101/2021.10.19.21265213

**Authors:** Garyfallos Konstantinoudis, Cosetta Minelli, Ana Maria Vicedo Cabrera, Joan Ballester, Antonio Gasparrini, Marta Blangiardo

## Abstract

**Background:** There is emerging evidence suggesting a link between ambient heat exposure and chronic obstructive pulmonary disease (COPD) hospitalisations. Individual and contextual characteristics can affect population vulnerabilities to COPD hospitalisation due to heat exposure. This study quantifies the effect of ambient heat on COPD hospitalisations and examines population vulnerabilities by age, sex, and contextual characteristics.

**Methods:** Individual data on COPD hospitalisation at high geographical resolution (postcodes) during 2007-2018 in England was retrieved from the small area health statistics unit. Maximum temperature at 1 km*×*1km resolution was available from the UK Met Office. We employed a case-cross over study design and fitted Bayesian conditional Poisson regression models. We adjusted for relative humidity and national holidays, and examined effect modification by age, sex, green space, average temperature, deprivation and urbanicity.

**Results:** After accounting for confounding, we found a 1.47% (95% Credible Interval 1.19% to 1.73%), increase in the hospitalisation risk for every 1°C increase in temperatures above 23.2°C (lags 0-2 days). We reported weak evidence of an effect modification by sex and age. We found a strong spatial determinant of the COPD hospitalisation risk due to heat exposure, that was alleviated when we accounted for contextual characteristics. 1 851 (95% CrI 1 576 to 2 079) COPD hospitalisations were associated with temperatures above 23.2°C annually.

**Conclusion:** Our study suggests that resources should be allocated to support the public health systems, for instance through developing or expanding heat-health alerts, to challenge the increasing future heat-related COPD hospitalisation burden.

**Key Messages:** *What is the key question?:* What is the short-term effect of heat exposure on COPD hospitalisation and which contextual/societal factors affect population vulnerability?

*What is the bottom line?:* For every 1°C increase in summer temperatures higher than 23.2°C, the risk of COPD hospitalisation increases by 1.47%, and populations in the North and in the South East are more vulnerable.

*Why read on?:* This large nationwide study in England using individual data quantifies the effect of heat exposure on COPD hospitalisations; these findings inform future policies regarding preparedness and resilience of public health systems against the increasing COPD burden due to the increasing temperatures.

## Introduction

Chronic obstructive pulmonary disease (COPD) is the most prevalent chronic respiratory disease worldwide, with point prevalence varying from 1.56% in Sub-Saharan Africa to 6.09% in Central Europe, eastern Europe, and central Asia in 2007.^1^ In England, COPD is a significant cause of morbidity and mortality, leading to 115 000 emergency admissions and 24 000 deaths per year.^2^ The causes of acute exacerbation of COPD are established and include factors such as sex, age, COPD severity and comorbidities.^3^ Environmental triggers of COPD hospitalisations such as air-pollution exposure have also been discussed extensively.^4^ There is emerging evidence suggesting a link between heat exposure and COPD hospitalisation, either directly or through exacerbating the effects of factors such as ozone concentration that are associated with these events.^5^

Several previous studies have examined the effect of high temperatures on COPD hospitalisations, reporting higher rates with heat exposure^6-8^ and heat waves.^9 10^ The majority of these studies are based on aggregated data (at the city or regional level),^6 9-11^ whereas only a few considered individual data.^7 8^ Use of individual data allows investigation of possible effect modification by individual factors such as age and sex, and it avoids ecological bias arising when group-level associations do not reflect associations at the individual level.^12^ Although previous studies have assessed the vulnerability related to individual factors, such as age and sex,^6 8^ contextual characteristics, such as green space, average temperature, deprivation and urbanicity, are still poorly characterised. Two of the previous studies have examined the spatial variation of the temperature effect on COPD hospitalisation, using however very coarse geographical resolution.^6 8^

In this nationwide study in England during 2007-2018, we investigated the effect of heat exposure on COPD hospital admissions using a semi-ecological framework. We took advantage of the individual data availability of the outcome and adopted a case-crossover study design that naturally accounts for time-constant variables at the individual patient level. Thus, we were able to account for factors like age, sex, comorbidities, deprivation as well as lifestyle characteristics such as physical activity through the study design. We also adjusted for time-varying confounders, such as air-pollution exposure and relative humidity and examined how the effect of temperature is modified by age, sex and in space. Last, we assessed the extent to which contextual characteristics, such as green space, deprivation, urbanicity and average temperature, contribute to the observed spatial variation of the effect of temperature.

## Methods

### Study population

We included inpatient hospital admissions from COPD in England during 2007-2018 as retrieved from Hospital Episode Statistics (HES) data held by the UK Small Area Health Statistics Unit, provided by the Health and Social Care Information Centre. Age, postcode of residence at time of the hospitalisation, and date of hospitalisation were available for each record. We focused only on admissions with acute exacerbation of COPD as primary diagnosis. We investigated the following diagnostic groups: J40–44 according to the International Classification of Disease version 10 (ICD10).^13^ The analysis is restricted to June, July and August.

### Exposure

Daily minimum and maximum temperatures were available at 1 km*×*1 km resolution from the UK Met Office with methods described elsewhere.^14^ In brief, the daily temperature in each grid-cell was estimated based on inverse-distance-weighted interpolation of monitoring data, also accounting for latitude and longitude, elevation, coastal influence, and proportion of urban land use. To assign daily temperature to health records, the postcode centroids of each patient were spatially linked to the 1km*×* 1km grid cell, applying a 100m fuzziness to the postcode location to fulfil governance requirements. We focused on daily maximum temperature, as we are interested in heat exposure, averaged over the day of hospitalisation and the preceding two days (lags 0-2 days) to estimate the cumulative health effects.^15-17^

### Covariates

We used hourly concentration of Ozone (O_3_) and atmospheric particulate matter that has a diameter of less than 2.5 *µm* (PM_2.5_), as retrieved from the unified model produced by the Met Office measured in *µg/m*^3. 18^ The model outcome is then post-processed to correct for bias using observational data. ^18^ For O_3_ we calculated the daily mean of the 8 hours of maximum O_3_, whereas for PM_2.5_ the daily mean concentration. The geographical resolution of the air-pollutants is 12 km*×*12 km for the years 2007-2011 and 2km*×*2km during 2012-2019. We adjusted for relative humidity (daily and at a 10 km*×*10 km grid) through a model that integrates MetOffice data on daily observations from the meteorological stations and monthly nationwide data as retrieved from HadUK,^14^ see Online Supplement Text S1.1. All covariates were included at lags 0-2 days, to match the exposure lags. O_3_, PM_2.5_ and relative humidity were included as linear terms in the model. We also accounted for the effect of national holidays through a dummy variable.

### Spatial effect modifiers

We selected these spatial effect modifiers based on consistency with the literature,^19^ data availability in England and a priori hypotheses, see Online Supplement Text S1.3. As a measure of green space we used the proportion of a region that is covered by green land such as woodland, agricultural land, grassland and other natural vegetated land as classified in the Land Cover Map 2015 (LCM15).^20^Deprivation is measured using the Index of Multiple Deprivation (IMD) 2015, as retrieved from the Ministry of Housing, Communities and Local Government.^21^ We used the quintiles of IMD in our analysis. For these two modifiers, we selected the year 2015 as the most representative data point, among the ones available, for our study period. Urbanicity (predominantly rural, urban with significant rural and predominantly urban) is based on the Office for National Statistics (ONS) classification in 2011 (the most recent year for which data was available at the time of analysis).^22^ We also incorporated the average temperature during 2007-2018, as a measure of adaptation on higher temperatures.^23^ Green space and average temperature were included as linear terms in the model. Due to power and computational considerations, all spatial effect modifiers were included at the lower tier local authority level (LTLA; Online Supplement Fig. S1).

### Statistical methods

We used a time-stratified case-crossover design, commonly used for analysing the effect of transient exposures.^24 25^ The temperature on the day of COPD hospitalisation (event day), is compared with the temperature on non-event days. In the case-crossover design a case serves as its own control, thus this design automatically controls for factors that do not vary or vary slowly over time, such as sex or deprivation. We selected non-event days on the same day of week and calendar month as the event day to avoid the overlap bias.^26^ Thus we could have maximum 4 non-event days per event day.

We modelled the effect of temperature on event compared to non-event days by specifying Bayesian hierarchical conditional Poisson models, with a fixed effect on the event/non-event day grouping.^19 27^ We accounted for recurrent hospitalisations by adding a random effect on each patient. For the main analysis, we treated relative humidity and national holidays as confounders and thus adjusted for them, but we did not adjust for air-pollutants because they were treated as mediators, see directed acyclic graph on Online Supplement Text S1.3.^28^ As the effect of temperature on health is typically non-linear,^19^ we used piecewise linear threshold models, to allow more flexible fits, but retain ease of interpretation. We considered nationwide thresholds, specified as the 50th, 55th …, 95th percentile of the daily temperatures. We selected the threshold based on the WAIC, an estimate of predictive accuracy, with smaller values indicating better fits.^29 30^ We then ran additional models allowing the effect of heat exposure (temperatures above the threshold) to vary by sex (male and female), age (0*-*64, 65*-*74, 75+) and space (LTLA). We additionally included the air-pollutants in these models to examine the sensitivity of the effect if the air-pollutants were confounders. For the spatial effect modification, we used the Besag-York-Mollie prior that assumes local dependency among adjacent LTLAs.^31^ We fitted this model with and without the spatial effect modifiers, while adjusting for confounders. The model is described in detail in the Online Supplement Text S1.2. Results are reported as medians and 95% Credible Intervals (CrI; 95% probability that the true values lies within this interval) of % increase in the hospitalisation risk for every 1°C increase in temperatures above the threshold;^32^ additionally, we report posterior probabilities of a positive % increase. For the spatially varying risk, we also reported posterior probabilities that the % hospitalisation risk is larger than the average % hospitalisation risk.

### Population attributable fraction

To calculate the population attributable fraction, we extended^33^ to incorporate the spatial dimension of the effect of heat exposure. We first calculated the cumulative heat exposure - COPD hospitalisation relative risk (RR_*s*_) for the *s*-th LTLA. We could then calculate the attributable fraction: AF_*s*_ = (RR_*s*_ *−* 1)*/*RR_*s*_. Let *n*_*s*_ be the number of hospitalisations at days above 23.2°C and *N*_*s*_ the total number of hospitalisations, then AF_*s*_(*n*_*s*_*/N*_*s*_) is the population attributable fraction, i.e., the number of COPD hospitalisations attributable to summer heat exposure. In our Bayesian formulation we were able to propagate all the random variable-specific uncertainty in our estimates.

### Sensitivity analyses

We repeated the main analysis for the lags 0, 1 and 2 independently. We also used b-splines to model the temperature effect and examined the linearity assumption above the threshold. All analyses are run in R-NIMBLE (Numerical Inference for Hierarchical Models Using Bayesian and Likelihood Estimation).^34^ The code for running the analysis is online available at https://github.com/gkonstantinoudis/COPDTempSVC.

## Results

### Population

We retrieved 1 570 288 COPD hospital records during 2007-2018 in England. After removing the duplicated records, the ones with place of residence outside England, the ones not occurred in summer months and the ones for which we could not sample non-event days, we had 320 411 records available for the analysis Fig. 1.

**Fig. 1.**
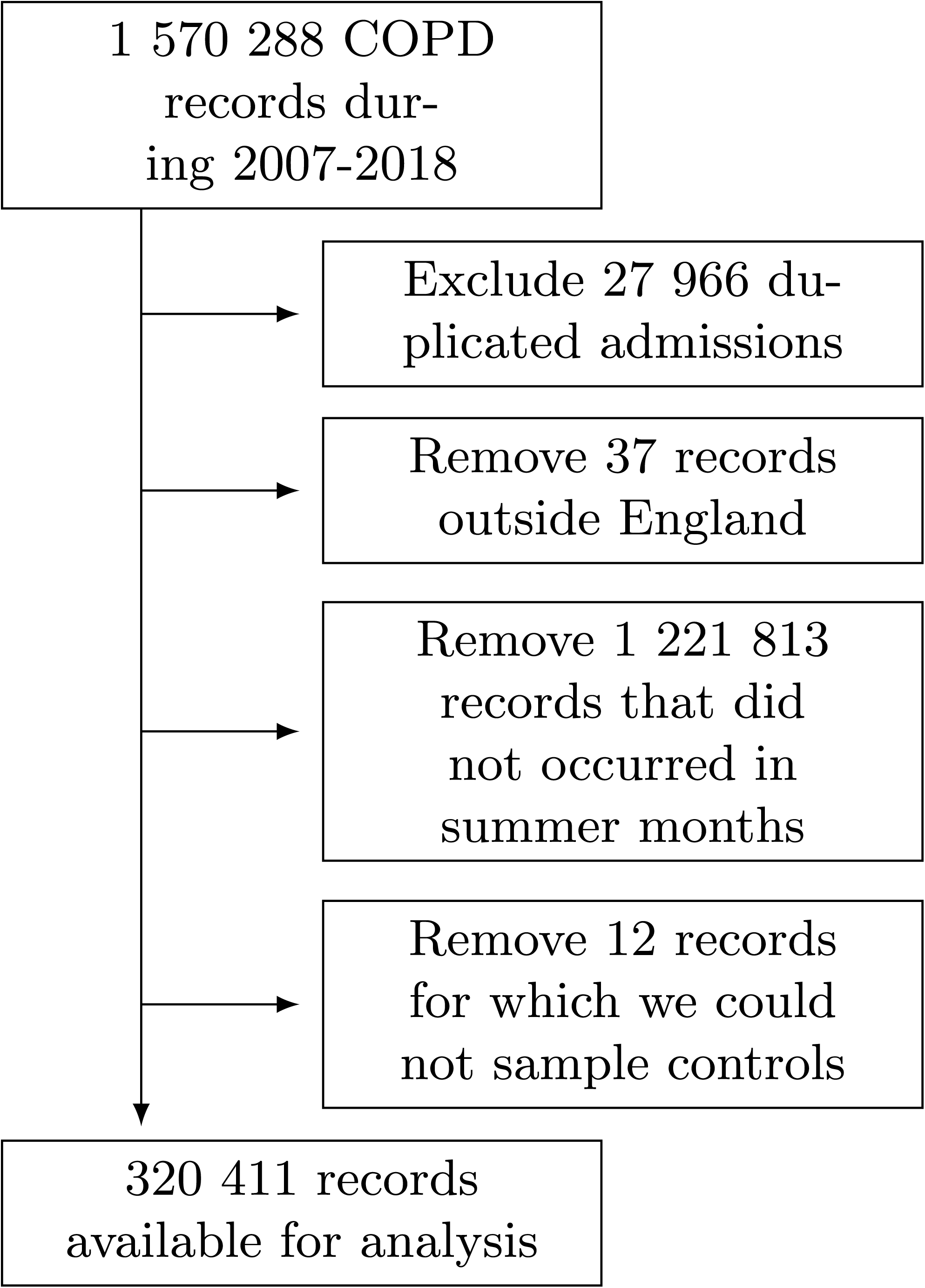
Flowchart of COPD hospitalisations.

### Exposure, Covariates and Effect modifiers

The median maximum temperature across England has increased from 19.42°C in 2007 to 22.20°C in 2018, Online Supplement Table S1. The median maximum temperature exposure is 20.91°C at lag 0 for event and 20.39°C non-event days, 20.97°C for event and 20.94°C for non-event days at lag 1, 20.92°C for event and 20.90°C for non-event days at lag 2 and 20.93°C for event and 20.92°C for non-event days at lag 0-2, Online Supplement Table S2. The distribution of the covariates across event and non-event days and the spatial distribution of the effect modifiers at the LTLA level can be found on the Online Supplement, Table S2-5, and Fig. S2-5.

### WAIC analysis

In the model adjusted for relative humidity and national holidays, the 80th percentile of the temperature (23.2°C) was the threshold minimising the WAIC, Online Supplement Table S6. We found a 0.37% (95% CrI 0.09% to 0.65%) increase in the COPD hospitalisation risk for every 1°C increase in temperatures below 23.2°C, Online Supplement Table S6. In contrast, the effect above 23.2°C was higher, namely 1.46% (95% CrI 1.19% to 1.71%), Online Supplement Table S6. All subsequent analyses were conducted using the 80-th percentile of the temperature as the threshold.

### Age and sex effect modification

In the unadjusted models, the percentage of risk increase in hospitalisations for every 1°C increase above the threshold varies from 0.92% (95% CrI 0.25% to 1.63%) in females 64 years old or younger to 1.56% (95% CrI 0.94% to 2.20%) in females aged 65-74, Fig. 2 and Online Supplement Table S6. After adjusting for relative humidity and national holidays the effects are slightly higher varying from 1.14% (95% CrI 0.39% to 1.84%) in females 64 years old or younger to 1.75% (95% CrI 1.13% to 2.41%) in males 65-74 years old, Fig. 2 and Online Supplement Table S7. Additionally adjusting for air-pollution substantially reduces the observed effect, Fig. 2 and Online Supplement Table S7.

**Fig. 2.**
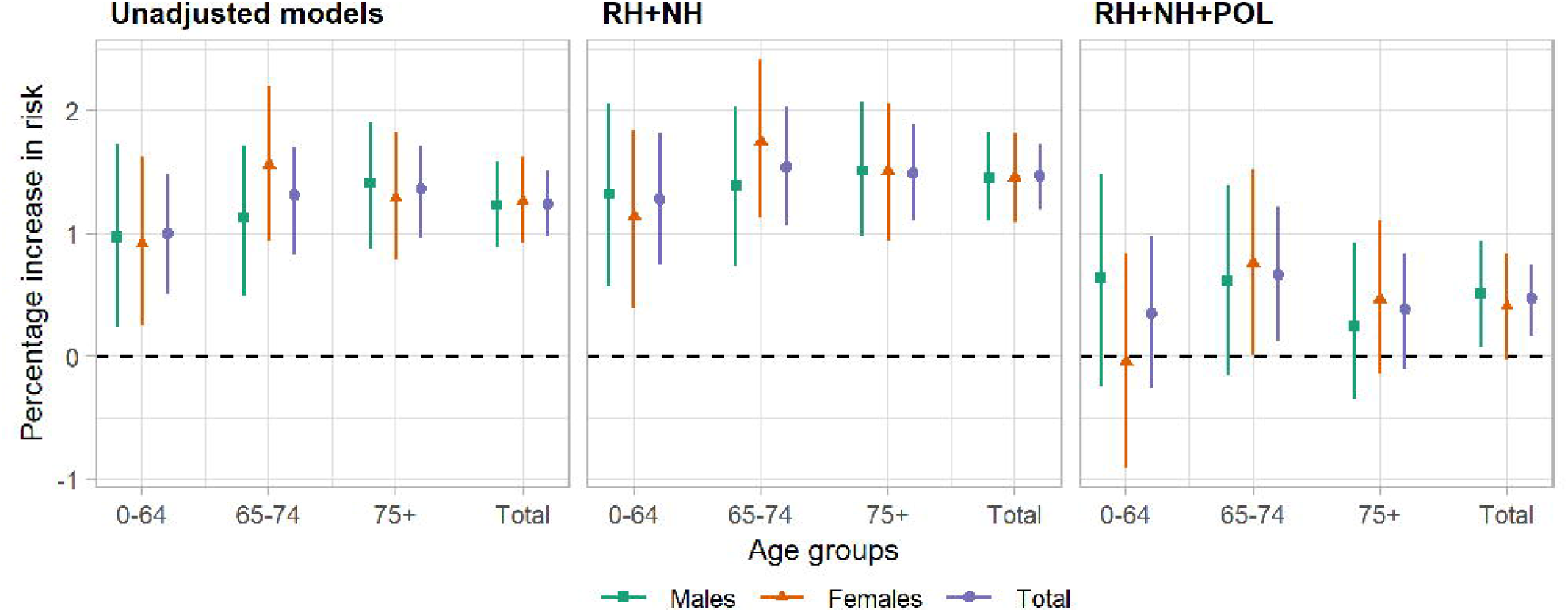
Percentage risk of COPD hospitalisation for every 1°C increase in the temperatures above 23.2°C during the summer months between 2007 and 2018, for the unadjusted (left panel), the model adjusted for relative humidity (RH) and national holidays (NL) (mid panel) and the model additional adjusted for air-pollution (POL) (right panel). Results are stratified by age (0-64, 65-74, 75+, total) and sex (male, female, total).

### Spatial effect modification

The spatial variation of the effect of heat exposure on COPD hospitalisations is shown on Fig. 3. The risk of COPD hospitalisation is less than 1.31% for every 1°C increase in heat exposure in South West, top left panel Fig. 3. In contrast, populations in the South East are more vulnerable: the probability that the effect of heat exposure is larger than the national average estimate ranges between 0.6 and 1, top right panel Fig. 3. After incorporating green space, deprivation, urbanicity, and average temperature, the observed variation of the effect of temperature is alleviated, bottom panels Fig. 3.

**Fig. 3.**
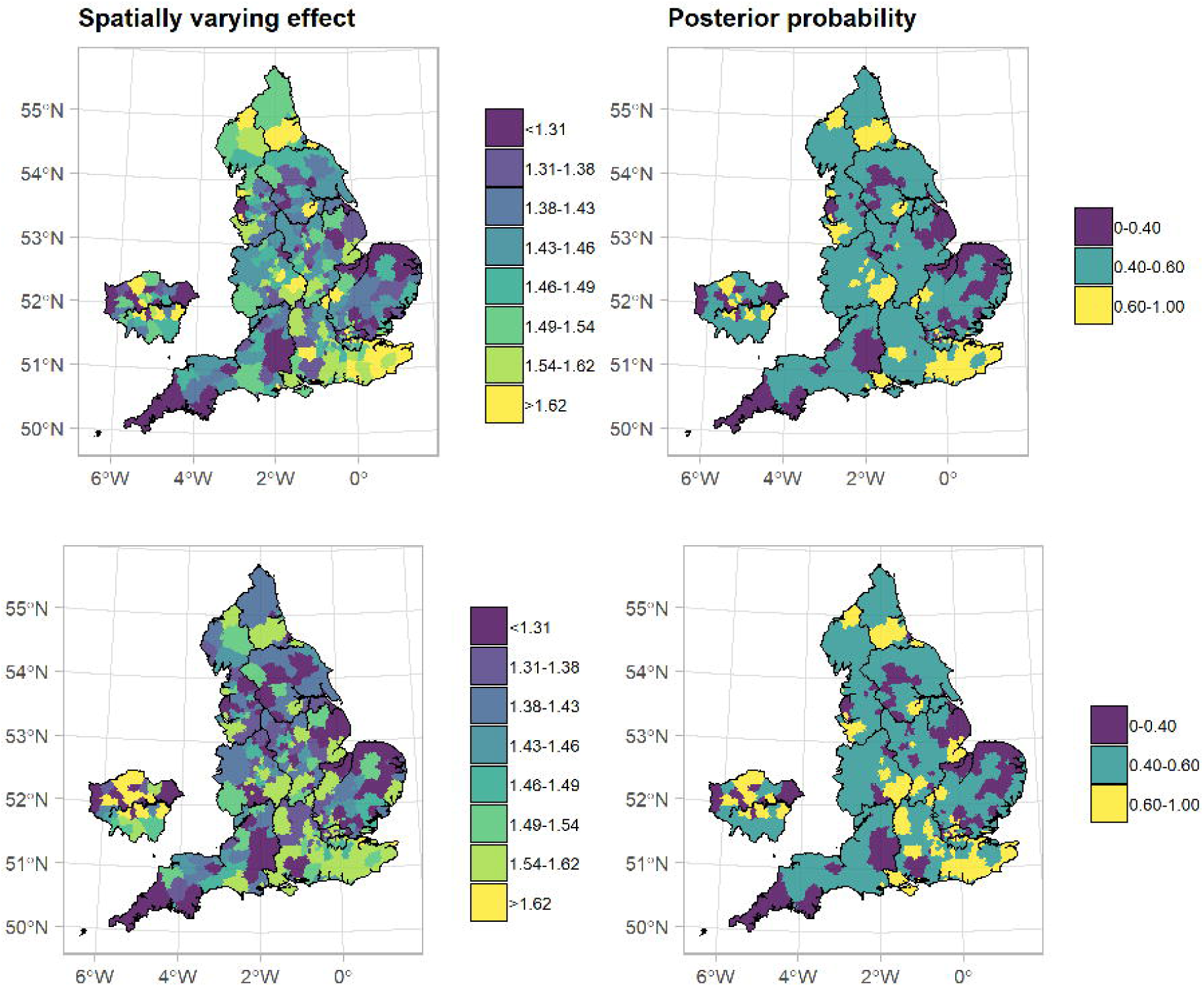
Median spatial COPD hospitalisation risk for every 1°C increase in the temperatures above 23.2°C and posterior probability that the risk is larger than the overall risk in England during the summer months between 2007 and 2018. The top panels refer to the model without incorporating contextual characteristics, whereas the panels below otherwise. All models were fully adjusted.

We found weak evidence that populations in areas with higher proportions of green space, larger average temperature and higher level of urbanicity are more resilient to COPD hospitalisations due to heat exposure, Table 1. If we increase an LTLA’s proportion of green space by 1%, the spatial effect of the heat exposure changes by -1.46% (95% CrI -6.99% to 4.39%), Table 1. For every 1°C increase in the average temperature per LTLA, the spatial effect of the heat exposure changes by -0.41% (95% CrI -1.49% to 0.71%), Table 1. The spatial effect of heat exposure in urban LTLAs with significant rural and predominantly urban LTLAs changes by -0.79% (95% CrI -3.10% to 1.51%) and -1.57% (95% CrI -4.16% to 0.96%) respectively compared with predominantly rural LTLAs, Table 1.

**Table 1.** Median,95% credible intervals of the percentage change of the heat exposure related spatial hospitalization risk due to green space, average temperature, index of multiple deprivation and urbanicity and probability that this percentage change is higher than 0.

| Effect modifier | Percentage change | Pr(% change > 0)<sup>1</sup> |
| --- | --- | --- |
| Green space<sup>2</sup> | -1.46 (-6.99 to 4.39) | 0.30 |
| Average temperature<sup>3</sup> | -0.41 (-1.49 to 0.71) | 0.22 |
| IMD<sup>4</sup> |  |  |
| Q1 | 1 |  |
| Q2 | 0.81 (-1.16 to 3.08) | 0.78 |
| Q3 | 1.57 (-0.76 to 4.06) | 0.91 |
| Q4 | 0.75 (-1.68 to 3.36) | 0.71 |
| Q5 | 1.62 (-1.31 to 4.49) | 0.85 |
| Predominantly Rural | 1 |  |
| Urban with significant rural | -0.79 (-3.10 to 1.51) | 0.25 |
| Predominantly urban | -1.57 (-4.16 to 0.96) | 0.12 |

<sup>1</sup> Posterior probability that the percentage change is larger than zero.
<sup>2</sup> Green space is the proportion of a region covered by green land such as woodland, agricultural land, grassland and other natural vegetated land.
<sup>3</sup> The average temperature is the mean summer temperature per LTLA during 2007-2018°C.
<sup>4</sup> Index of multiple deprivation. IMD is calculated based on the following domains: a. income, b. employment, c. education, skills and training, d. health and disability, e. crime, f. barriers to
housing and services and g. living environment deprivation. Q1 denotes the most deprived areas, whereas Q5 the least deprived.

### Population attributable burden

We found that 1 851 (95% CrI 1 576 to 2 079) COPD hospitalisations were associated with temperatures above 23.2°C annually. This accounts for 7.8% (95% CrI 6.7% to 8.8%) of the total COPD hospitalisations during the summer months from 2007 to 2019. The proportion of COPD hospitalisations attributable to temperatures above the threshold has a clear spatial structure and is more than 8% in East Midlands, East of England, London and South East, while it is below 5% in the South West (Fig. 4).

**Fig. 4.**
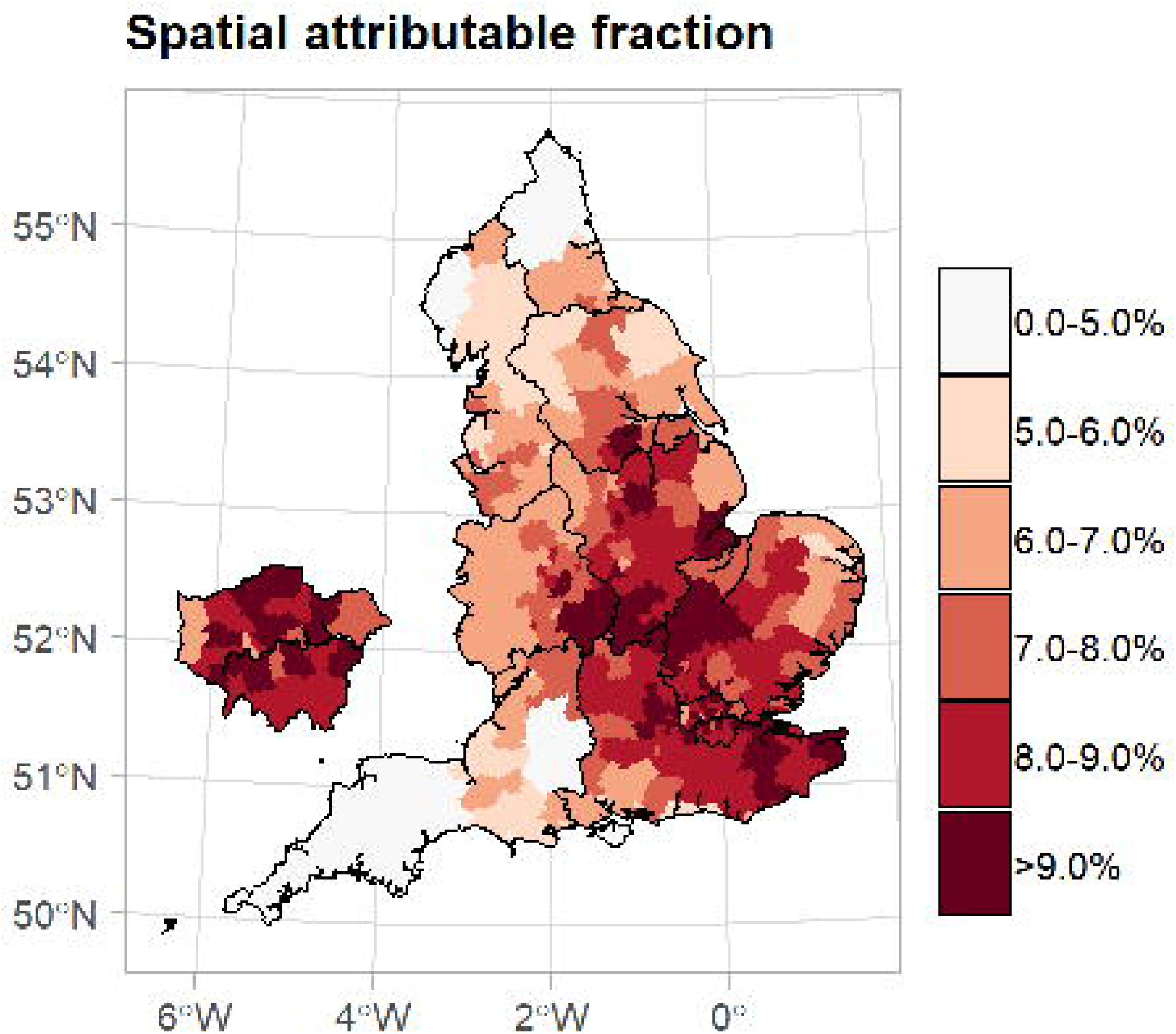
The percentage of COPD hospitalisations by lower tier local authorities attributed to exposure to summer temperatures above 23.2°C during 2007-2018 in England. This effect assumes a causal relationship between heat exposure and COPD hospitalisation risk. The island on the left is a zoomed version of London.

### Sensitivity analysis

The lag with the highest influence was lag 1 with the risk of COPD hospitalisation being 1.37% (95% CrI 1.14% to 1.58%) for every 1°C increase in heat exposure. For lag 0 and lag 2 the point estimate was still positive, but lower in magnitude, 0.71% (95% CrI 0.50% to 0.93%) and 1.01% (95% CrI 0.78% to 1.24%) respectively, likely due to the correlation with temperatures at lag 1. The linearity assumption above the 23.2°C threshold looks reasonable, Online Supplement Fig. S6.

## Discussion

This is the first nationwide case-crossover study in England investigating the short-term effects of heat exposure on COPD hospitalisation. After accounting for confounding, the results indicate that for every 1°C increase in heat exposure the COPD hospitalisation risk increases by 1.47% (95% CrI 1.19% to 1.73%), with evidence that PM_2.5_ and O_3_ mediate this relationship. We found weak evidence of an effect modification by sex and age. The attributable burden of heat exposure has a clear spatial structure, with areas in East Midlands, East of England, London and South East affected the most. Assuming a causal relationship, 7.8% (95% CrI 6.7% to 8.8%) of COPD hospitalisations could be attributed to heat exposure during the summer months between 2007 and 2018.

The main strength of our study is the availability of postal codes, exploiting the highest spatial resolution available for linkage with the exposure and confounding factors. Such geographical resolution is expected to minimise misclassification resulting from any spatial misalignment between the outcome and exposure/confounder. The availability of individual data for the outcome also minimises ecological bias,^12^ while guaranteeing high statistical power due to the population-based nature of the study. We ascertained hospital records from NHS digital covering almost all hospitalisation occurred in the public sector in England during 2007-2018. Our study has some limitations. First, residential temperature does not reflect the actual temperature exposure of an individual, as individuals are exposed to different temperatures in the course of the day. In addition to this, the outdoor temperature, as provided by Met Office does not reflect the actual temperature exposure inside the house. Nevertheless, in line with most of the studies in this field and given the lack of more precise individual exposure data, we used residential temperature outdoors as a proxy for the individual exposure. To allow for flexible fits, we used a linear threshold model. More complex relationships may need multiple thresholds, however the WAIC analysis suggested that the linearity assumption suffices. Although we adjusted for the main COPD hospitalisation environmental contributors, we could not evaluate other potential confounders (e.g., seasonal allergies and pollen counts) due to the lack of available data. Additionally, exposure to other air-pollutants, such as NO_2_, SO_2_, might also confound the observed relationship; we decided to adjust for PM_2.5_ and O_3_ as they seem to have a larger impact on COPD hospitalisation and to avoid potential collinearity with other pollutants.

Our results can be compared with studies examining COPD hospital admissions and ambient temperatures during the hottest months.^6 11 35 36^ Our study is in line with a US study including 12.5 million participants that found a 4.7% (95% CrI 3.9 to 5.5%) increase in the COPD hospitalisation rate at lag 0 for every 5.6°C increase in the average daily temperature during May-September.^6^ Our study is also in line with a case-crossover study in Brazil that reported a 5% (95% CrI 4% to 6%) increase in the hospitalisation odds for every 5°C increase in the average temperature (0-3 lags) during the 4 hottest months.^11^ In contrast, a study in New York reported a 7.64% increase in the risk of COPD admissions for each 1°C increase in daily mean apparent temperature above 32°C.^36^ A study in 12 European cities, reported a 4.5% (95% CrI 1.9 to 7.3) and 3.1% (95% CrI 0.8 to 5.5) increase in total respiratory admissions (the majority being COPD) in Mediterranean and North-Continental cities, respectively, for every for each 1°C increase in the maximum apparent temperature (lag 0-3 days) above the 90th percentile.^37^ A study in Taiwan reported negative correlation between the average daily temperature and emergency admissions with COPD, but a 14% increase in the emergency COPD admissions when the diurnal temperature range is larger than 9.6°C.^35^

We found weak evidence of an effect modification by age and sex, but discrepancies in vulnerability in space. A previous study in Brazil, reported higher COPD hospitalization odds for women and the older people.^11^ In the models adjusted for relative humidity and national holidays, in line with a previous study in the US,^6^ the age group 65-74 was the most vulnerable. Some spatial variability by regions or counties was also observed in previous studies in Brazil and the US, potentially due to socioeconomic characteristics or exposure to higher average summer temperatures.^6 11^ In our study, green space, average temperature, deprivation and urbanicity explained some of the observed variation in the observed spatial vulnerabilities, the evidence of an effect was however inconclusive.

Some discrepancies of our results compared with previous studies can have multiple explanations. Previous studies reporting higher effect estimates had available coarser geographical resolution (city or county level), leading to inadequate adjustment for confounding, as confounders, can vary in high geographical resolution.^11 35 37 38^ Differences in the definition of the outcome can also lead to the discrepancies as previous studies have used the apparent temperature,^36 37^ or diurnal temperature range,^35^ while others, more in line with our approach, the daily mean.^6 11^ Decisions regarding the selection of the temperature threshold, the warm-season months and the lags can also partly explain the observed difference in the effect estimates. Most previous studies adjusted for air-pollution,^6 36 37^ while we did not, as we assumed that air-pollution is a mediator;^28^ when we added air-pollution to model the effect of heat exposure was much reduced.

Acute COPD episodes are associated with airways and systemic inflammation but also with cardiovascular comorbidity and may be triggered by exposures to heat.^37^ Exposure to ambient heat can lead to heat dissipation through hyperventilation and may trigger dynamic hyperinflation and dyspnoea in patients with pre-existing COPD.^6 11^ The higher risk of COPD hospitalisation in the 65-74 age group observed in our study could be explained by the inability of this frail population to dissipate excess heat through circulatory adjustment, and exposure to extreme temperatures increases their risk of developing pulmonary vascular resistance secondary to peripheral pooling of blood or hypovolemia.^37^ In addition, older populations are of higher risk to have cardiovascular comorbidities, which are hypothesised to increase the risk of COPD hospitalisations associated with heat exposure. Nevertheless, such evidence is inconclusive.^37^ We also reported a weak protective effect of higher average temperatures, arguing towards protective adaptation to heat, possibly related to differences in housing stock or behaviour during hot weather.^11^ We observed weak evidence of increased resilience in populations in more deprived areas and in areas with higher degrees of urbanicity. Although this evidence is inconclusive, potential factors that could confound the observed effect include differences in demographics, for instance ethnicity.

Previous studies examining future trends in COPD, population demographics and temperature changes have predicted a higher COPD prevalence, a raise in the average age of the population and increased global temperatures.^39-41^ Resources should be allocated to support the preparedness and resilience of public health systems, for instance through developing or expanding heat-health alerts, to challenge the increasing heat exposure related COPD hospitalisation burden.

## Data Availability

SAHSU does not have permission to supply data to third parties. For reproducibility purposes we have simulated data and provided the code used for the analysis in https://github.com/gkonstantinoudis/COPDTempSVC.

## Acknowledgements

We thank Hima Daby, Gajanan Natu, Eric Johnson and Bethan Davies for their help with data acquisition, storage, preparation, and governance. All authors acknowledge infrastructure support for the Department of Epidemiology and Biostatistics provided by the NIHR Imperial Biomedical Research Centre (BRC). Hospital Episode Statistics data are copyright © 2021, re-used with the permission of NHS Digital. All rights reserved. The Hospital Episode Statistics data were obtained from NHS Digital.

## Contributors

G.K and M.B. conceived the study. M.B. supervised the study. G.K. developed the initial study protocol and discussed it with M.B., C.M., A.M.V.C., J.B and A.G.. G.K. developed the statistical model, prepared the covariate data and led the acquisition of hospitalisation data. M.B. validated the code. G.K. ran all the analysis and wrote the initial draft. All the authors contributed in modifying the paper and critically interpreting the results. All authors read and approved the final version for publication.

## Patient consent for publication

Not required.

## Ethics

The analyses were covered by national research ethics approval from the London-South East Research Ethics Committee (Reference 17/LO/0846). Data access was covered by the Health Research Authority Confidentiality Advisory Group under section 251 of the National Health Service Act 2006 and the Health Service (Control of Patient Information) Regulations 2002 (Reference 20/CAG/0028).

