## Supplementary material for "Ambient heat exposure and COPD hospitalisations in England: A nationwide case-crossover study during 2007-2018": Online Supplement

### S1 Text

#### S1.1 Modelling relative humidity

##### S1.1.1 Model

Data on relative humidity in England during 1862-2019 is available nationwide from MetOffice through the HadUK-Grid product (`https://catalogue.ceda.ac.uk/`). The spatial resolution of HadUK-Grid can vary from 1km×1km to 60km×60km, nevertheless the highest temporal resolution available are months. MetOffice also provides daily data on relative humidity during 1853-2019 for each meteorological station through the Met Office Integrated Data Archive System (MIDAS) product (`https://catalogue.ceda.ac.uk/`). To retrieve daily relative humidity data nationwide and not only at the meteorological stations, we employ the following modelling framework:

Let $Y_{jkt}(s)$ be the arcsin transformation of the relative humidity from MIDAS and $X_{kt}(s)$ the nationwide relative humidity from HadUK-Grid in location $s$, day $j$, month $k$ and year $t$:

$$\begin{aligned}
Y_{jkt}(s) &\sim \text{Normal}\big(\mu_{jkt}(s), \sigma_1\big) \\
\mu_{jkt}(s) &= \beta_0 + bX_{kt}(s) + \gamma_j + \omega_t + u(s) \\
\gamma_j &\sim \text{AR1}(\sigma_2, \rho) \\
\omega_t &\sim \text{Normal}(0, \sigma_3^2) \\
u(s) &\sim \text{GMRF}(\sigma_4, \phi) \\
\sigma_1, \sigma_2, \sigma_3, \phi, \rho &\sim \text{PCpriors} \\
\beta_0, b &\sim N(0, \delta)
\end{aligned} \tag{1}$$

wheres $\mu_{jkt}(s)$ and $\sigma_1$ be the variables of the normal distribution, $\beta_0$ and $b$ the regression coefficients, $\gamma_j$ a random effect capturing the daily temporal trends, $\omega_t$ a random effect capturing the yearly trends, $u(s)$ the spatial autocorrelation term (based on the Stochastic Partial Differential Equation Approach [1]) and $\sigma_2, \sigma_3, \sigma_4, \rho,$ and $\phi$ the corresponding variance and correlation hyperparameters.

The specification of the PCpriors of the hyperparameter is the following: for $\sigma_1$ we specify that the probability of observing arcsin relative humidity larger than 10 is 0.10. Similarly for $\sigma_2$ and $\sigma_3$, we specify that the probability of observing arcsin relative humidity larger than 1 due to the temporal trends is 0.10. For the correlation parameter $\rho$ of the autoregressive process of order 1, we select a probability of 0.5 for correlations of 0.5 reflecting our lack of knowledge with respect to the correlation structure in the data. For the Gauss Markov Random Field

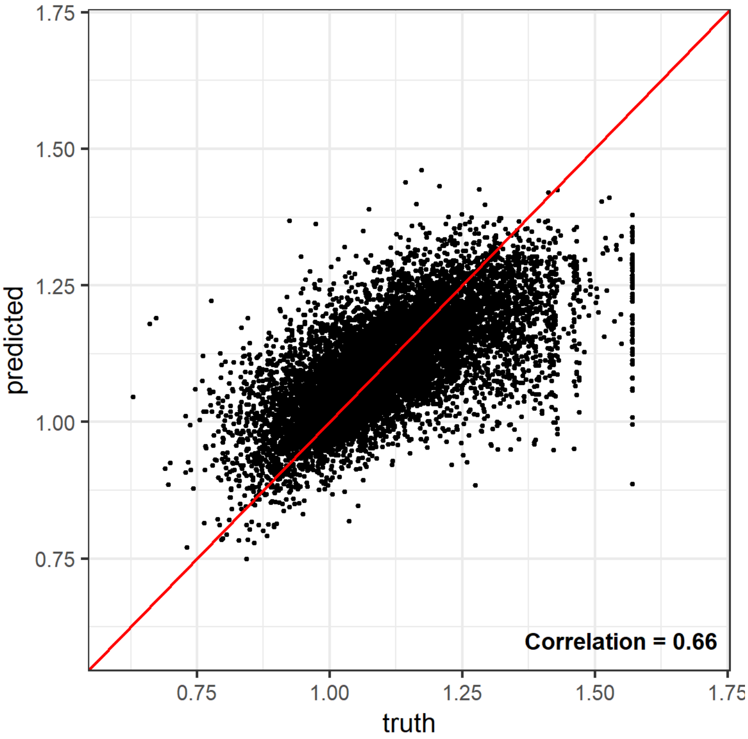

Figure 1.1. Predicted versus true values of the arcsin relative humidity for a sample of 10,000 values in the summer months during 2007-2018 in England.

(GMRF) term we select the standard deviation hyperparameter $\sigma_4$ as in the temporal case, whereas for the range parameter $\phi$ we select ranges larger that 10km with probability of 0.5. For more information about the PCpriors and their mathematical formulation see [2, 3]. Lastly, $\delta$ was fixed to 0.001 for the intercept, whereas to 0.1 for b.

###### S1.1.2 Cross-validation

We performed the following leave one out cross validation scheme: Let N be the total number of meteorological stations during 2007-2019, first we divided N randomly by 10 groups, and for each (out of the 10) step we excluded the entire time series of the group of the randomly sampled N/10 meteorological stations. Figure 1.1 shows the results of the cross validation, and in particular a scatterplot between the observed and predicted values. The correlation between truth and predicted is relatively high, ie 0.66, indicating that our model have good predictive ability.

###### S1.1.3 Results

Table 1.1 shows the results of Model 1, Figure 1.3 shows the mean of the daily median of relative humidity in 2013 for the 3 summer months. The maps show that areas around London had lower relative humidity during the summer months in 2013. The relative humidity seems to be consistently higher in South West during the

Table 1.1. Mean, standard deviation, median and 94% credible intervals of the intercept, the covariate and the hyperparameters of Model 1.

| Random variables | mean | sd | median | 2.5% | 97.5% |
|---|---|---|---|---|---|
| $\beta_0$ | 0.144 | 0.009 | 0.144 | 0.126 | 0.161 |
| b | 0.012 | 0.000 | 0.012 | 0.012 | 0.012 |
| $1/\sigma_1^2$ | 101 | 0.285 | 101 | 101 | 102 |
| $1/\sigma_2^2$ | 172 | 8.30 | 172.26 | 156.30 | 189 |
| $\rho$ | 0.386 | 0.028 | 0.384 | 0.332 | 0.440 |
| $1/\sigma_3^2$ | 68.1 | 26.3 | 63.8 | 29.8 | 131 |
| $1/\sigma_4^2$ | 0.063 | 0.007 | 0.063 | 0.052 | 0.079 |
| $\phi$ | 8,384 | 1,557 | 8,352 | 5,493 | 11,600 |

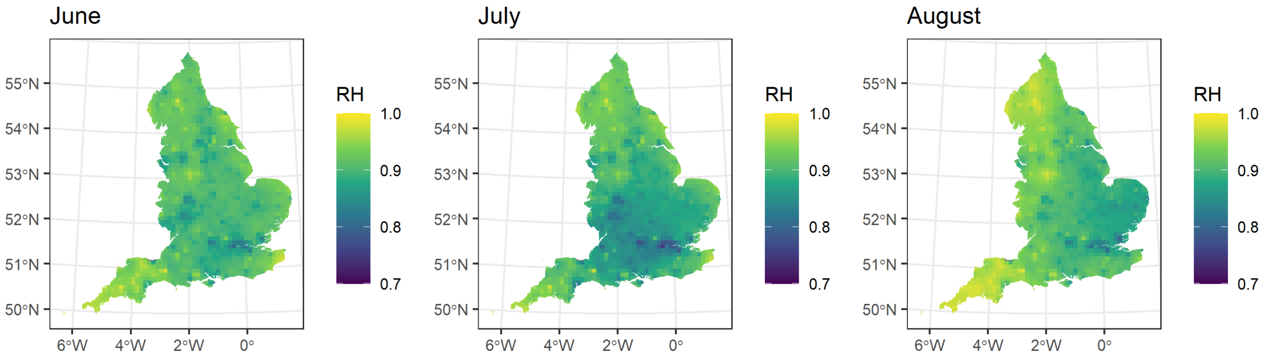

Figure 1.3. Maps of mean of median relative humidity by summer month in 2013.

summer 2013.

#### S1.2 Statistical analysis

In this subsection we will introduce the mathematical notation of the models used in the main analysis.

##### S1.2.1 WAIC analysis

Let $Y_{tjk}$ be the case-control identifier for the chronic obstructive pulmonary disease (COPD) hospitalisation for the event (case or control) at time $t$, in the $j$-th case-control group and $k$-th patient. Let also $X_1t$ be the temperature at $t$ event $Z_{1t}, Z_{2t}$) a vector denoting the different confounders (relative humidity and holiday) at the $t$-th time point. Then:

$$\begin{aligned}
Y_{tjk} &\sim \text{Poisson}(\mu_{itjk}) \\
\log(\mu_{tjk}) &= \alpha_1 I(X_{1t} < c_l)X_{1t} + \alpha_2 I(X_{1t} \geq c_l)X_{1t} + \sum_{m=1}^{2} \beta_m Z_{mt} + u_j + w_k \\
u_j &\sim N(0, 100) \\
w_k &\sim N(0, \sigma_1^2) \\
a_1, a_2, \beta_1, \dots \beta_4 &\sim N(0, 1) \\
\sigma_1 &\sim \text{Gamma}(1, 2)
\end{aligned}$$

In the above equation, $a_1$ is the effect of temperatures lower than the threshold $c$, $a_2$ is the effect of temperatures higher or equal than the threshold $c$, $I(\cdot)$ an indicator function, $\beta_1$, $\beta_2$ the effects of the confounding, $u_j$ a fixed effect on the $j$-th case control group and $w_k$ a random effect to account for recurrent hospitalisations. The normal distributions read N(mean, variance). We ran the above model for the different temperature thresholds $c_l$ for $l = 1, 2, \dots, 10$ representing the 50-th, 55-th, ... 95-th percentiles of the temperature and computed the WAIC [4]. Removing the term $\sum_{m=1}^{2} \beta_m Z_{mi}$ and the corresponding priors of $\beta_1, \beta_2$ results in the unadjusted models.

##### S1.2.2 Age-sex effect modification

Let $c_*$ be the temperature threshold that minimises the WAIC from Step 1. Expanding the indices of the above model results in the models for the age and sex effect modification. Let $g$ be the age-sex index representing individuals aged $0 - 64$, $65 - 74$ and $> 75$ years old or the total group and males, females or the total group.

The above model can be rewritten as follows:

$$
\begin{aligned}
Y_{tjkg} &\sim \text{Poisson}(\mu_{tjkg}) \\
\log(\mu_{tjkg}) &= \alpha_1 I(X_{1tg} < c_*)X_{1tg} + \alpha_2 I(X_{1tg} \geq c_*)X_{1tg} + \sum_{m=1}^{2} \beta_m Z_{mtg} + u_j + w_k \\
u_j &\sim N(0, 100) \\
w_k &\sim N(0, \sigma_1^2) \\
a_1, a_2, \beta_1, \dots \beta_5 &\sim N(0, 1) \\
\sigma_1 &\sim \text{Gamma}(1, 2)
\end{aligned}
$$

##### S1.2.3 Spatial effect modification

On the third step of the analysis we let the coefficient of the temperature higher than $c_*$ vary by lower tier local authorities (LTLA). Let $H_{1i}, \dots, H_{8i}$) be the spatial effect modifiers representing the green space, the quintiles of deprivation, the urbanicity categories and the average temperature in the $s$-th LTLA. We can write:

$$
\begin{aligned}
Y_{tjk} &\sim \text{Poisson}(\mu_{tjk}) \\
\log(\mu_{tjk}) &= \alpha_1 I(X_{1t} < c_*)X_{1t} + \alpha_{2s} I(X_{1t} \geq c_*)X_{1t} + \sum_{m=1}^{2} \beta_m Z_{mt} + u_j + w_k \\
\alpha_{2s} &= \alpha_2 + \sum_{q=1}^{8} \gamma_q H_{sq} + v_s + b_s \\
w_k &\sim N(0, \sigma_1^2) \\
v_s &\sim N(0, \sigma_2^2) \\
b_s | b_{-s} &\sim N\left( \frac{\sum_{s\sim r} w_{rs} b_s}{\sum_{s\sim r} w_{rs}}, \frac{\sigma_3^2}{\sum_{s\sim r} w_{rs}} \right) \\
u_j &\sim N(0, 100) \\
a_1, \beta_1, \dots \beta_4, \gamma_1, \dots, \gamma_8 &\sim N(0, 1) \\
a_2 &\sim N(0.0425, 0.0039^2) \\
\sigma_1, \sigma_2, \sigma_3 &\sim \text{Gamma}(1, 2).
\end{aligned}
$$

$w_{rs}$ are neighborhood weights and are 1 when the $r$ and $s$ LTLAs are neighboring (we write $s \sim r$) and 0 otherwise, $\gamma_1, \dots, \gamma_8$ are the effects of the spatial effect modifiers and the $v_s + b_s$ the BYM prior [5]. Unstructured overdispersion is captured on $v_s$ and spatial autocorrelation on $b_s$. The hyperparameters $\sigma_2^2, \sigma_3^2$ are the variance parameters of the unstructured and structured random effects. Removing the term $\sum_{q=1}^{8} \gamma_q H_{sq}$ and the corresponding priors of $\gamma_1, \ldots, \gamma_8$ results in the model without the adjustment for spatial effect modifiers, while allowing the effect of warm temperatures to vary in space.

#### S1.3 Confounders/ Mediators/ Effect modifiers

Directed acyclic graph for the relationship between temperature and hospitalisations for chronic obstructive pulmonary disease (COPD).

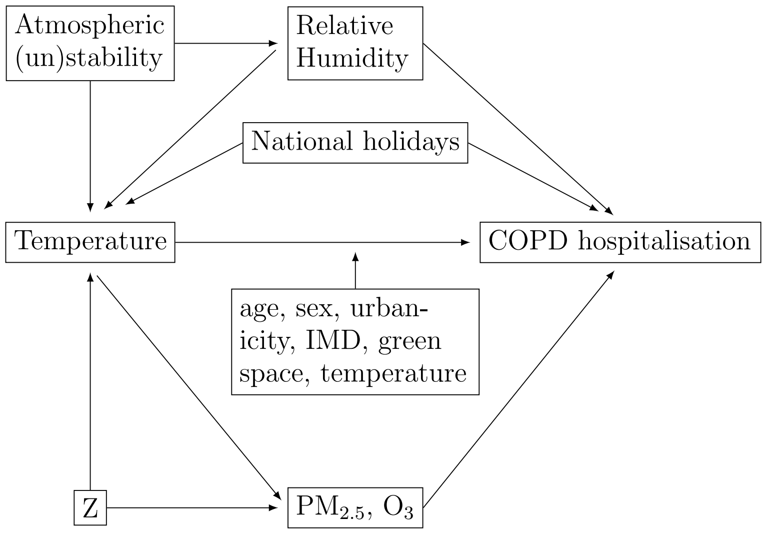

- **Relative humidity**: Previous studies has reported an association between relative humidity and COPD hospitalisations [6]. Relative humidity and temperature are both affected by factors such as atmospheric (un)stability and climate dynamics. Nevertheless, (soil and air) humidity determines the fraction of radiation (coming from the Sun and absorbed mainly by the surface) that is transformed into latent and sensible heat. Latent heat is generated due to phase transition of water, e.g., evaporation. The remaining radiation is transformed into sensible heat, leading to temperature changes [7]. Thus, relative humidity is likely a confounder.

- **National holidays**: can affect individual behaviours with respect to seeking health care services but also through other behaviours that can affect the temperature. Thus, holidays can be a potential confounder [8].

- **Air-pollution**: Previous studies have reported an association between short term exposure to $PM_{2.5}$ and $O_3$, and COPD hospitalisations [9, 10]. Thus these air-pollutants are expected to be correlated with COPD hospitalisations. Although temperature and air-pollutants have their own causal factors, e.g. air pollution emissions, they are also likely to have a shared cause Z, an example could be the already mentioned atmospheric (un)stability and climate dynamics, and temperature to affect $PM_{2.5}$ and $O_3$ concentration [11]. Thus, $PM_{2.5}$ and $O_3$ are likely to be mediators.

- **Effect modifiers**: The effect of temperature on COPD hospitalisations can be modified by, among other factors, age, sex, urbanicity, green space and average temperature. We selected these effect modifiers based on 1. consistency with the literature [8] and 2. Clear hypotheses about the mediation: We included age as the elderly have been reported to be more vulnerable, sex as differences can arise due to different lifestyle, occupational or biological factors, averaged temperature to account for potential adaptation to higher temperatures, urbanicity, to examine if urban heat island modifies the effect of temperature, and green space. Green space may reduce health risks in urban populations by removing air pollution, reducing noise, cooling temperature, enhancing physical activities, reducing psychological stress, and interaction with a clean environment [12]. 3. As the main interest of the current analysis was spatial effect modification, we did not include factors that vary significantly in time, such as air-pollution.

Table S1: Mean, standard deviation (sd), median, interquartile range (IQR), min and max of maximum summer temperature [$^o$C] across England during 2007-2018.

| year | mean | sd | median | IQR | min | max |
|---|---|---|---|---|---|---|
| 2007 | 19.39 | 2.71 | 19.42 | 3.31 | 4.00 | 30.31 |
| 2008 | 19.63 | 2.90 | 19.39 | 3.47 | 6.35 | 30.20 |
| 2009 | 20.26 | 3.28 | 20.27 | 4.17 | 2.25 | 31.97 |
| 2010 | 20.49 | 3.25 | 20.38 | 4.28 | 6.41 | 32.83 |
| 2011 | 19.38 | 3.10 | 19.15 | 3.98 | 5.32 | 33.41 |
| 2012 | 19.05 | 3.42 | 18.89 | 4.27 | 2.83 | 33.05 |
| 2013 | 21.10 | 3.78 | 20.88 | 5.37 | 5.76 | 34.09 |
| 2014 | 20.63 | 3.16 | 20.53 | 4.33 | 6.38 | 32.30 |
| 2015 | 19.87 | 3.37 | 19.79 | 4.23 | 3.03 | 36.67 |
| 2016 | 20.46 | 3.36 | 20.31 | 4.09 | 5.26 | 34.21 |
| 2017 | 20.37 | 3.45 | 20.17 | 4.09 | 5.49 | 34.48 |
| 2018 | 22.42 | 3.98 | 22.20 | 5.86 | 4.98 | 35.66 |

Table S2: Median and interquartile range (IQR) of the temperature [°C], daily mean $PM_{2.5}$ [$\mu g/m^3$], daily mean of the 8 hours of maximum $O_3$ [$\mu g/m^3$] and relative humidity [%] across event, non-event days and the different lags preceding the hospitalisation event.

| | | Event days | | Non-event days | |
|---|---|---|---|---|---|
| Covariate | lag | Median | IQR | Median | IQR |
| Temperature | 0 | 20.91 | 4.17 | 20.93 | 4.13 |
| | 1 | 20.97 | 4.21 | 20.94 | 4.13 |
| | 2 | 20.92 | 4.23 | 20.90 | 4.15 |
| | 0-2 | 20.93 | 3.73 | 20.92 | 3.67 |
| $PM_{2.5}$ | 0 | 9.23 | 5.60 | 9.10 | 5.45 |
| | 1 | 9.25 | 5.57 | 9.07 | 5.38 |
| | 2 | 9.23 | 5.44 | 9.06 | 5.25 |
| | 0-2 | 9.24 | 4.54 | 9.08 | 4.38 |
| $O_3$ | 0 | 65.5 | 22.37 | 65.00 | 21.91 |
| | 1 | 66.08 | 22.68 | 65.37 | 21.82 |
| | 2 | 66.21 | 22.35 | 65.58 | 21.78 |
| | 0-2 | 65.94 | 19.45 | 65.31 | 18.81 |
| Relative humidity | 0 | 0.90 | 0.11 | 0.90 | 0.11 |
| | 1 | 0.90 | 0.11 | 0.90 | 0.11 |
| | 2 | 0.90 | 0.11 | 0.90 | 0.11 |
| | 0-2 | 0.90 | 0.08 | 0.90 | 0.08 |

Table S3: Mean, standard deviation (sd), median, interquartile range (IQR), min and max of daily mean $PM_{2.5}$ [$\mu g/m^3$] exposure across England during summers 2007-2018.

| year | mean | sd | median | IQR | min | max |
|---|---|---|---|---|---|---|
| 2007 | 8.71 | 5.51 | 7.47 | 5.13 | 0.10 | 76.97 |
| 2008 | 8.71 | 6.07 | 7.37 | 4.17 | 0.00 | 69.92 |
| 2009 | 8.81 | 4.55 | 7.48 | 4.14 | 1.15 | 62.24 |
| 2010 | 9.19 | 5.03 | 7.97 | 4.78 | 0.70 | 52.42 |
| 2011 | 9.20 | 4.50 | 8.03 | 4.41 | 0.87 | 71.39 |
| 2012 | 8.28 | 4.97 | 7.04 | 4.78 | 0.35 | 84.33 |
| 2013 | 10.42 | 7.12 | 8.20 | 8.26 | 0.00 | 75.41 |
| 2014 | 8.58 | 4.83 | 7.39 | 5.43 | 0.31 | 53.73 |
| 2015 | 7.86 | 4.88 | 6.75 | 5.79 | 0.03 | 44.60 |
| 2016 | 8.60 | 8.20 | 6.07 | 6.51 | 0.00 | 73.65 |
| 2017 | 8.05 | 6.04 | 6.20 | 6.20 | 0.00 | 65.88 |
| 2018 | 8.67 | 5.67 | 7.13 | 6.07 | 0.00 | 52.24 |

Table S4: Mean, standard deviation (sd), median, interquartile range (IQR), min and max of daily mean of the 8 hours of maximum $O_3$ $[\mu g/m^3]$ exposure across England during summers 2007-2018.

| year | mean | sd | median | IQR | min | max |
|---|---|---|---|---|---|---|
| 2007 | 70.15 | 18.75 | 67.33 | 18.59 | 5.99 | 198.68 |
| 2008 | 71.32 | 18.91 | 69.35 | 21.86 | 0.19 | 166.95 |
| 2009 | 69.10 | 20.51 | 64.09 | 22.77 | 3.09 | 180.97 |
| 2010 | 68.11 | 19.87 | 64.57 | 23.99 | 12.07 | 157.44 |
| 2011 | 67.02 | 16.06 | 65.18 | 18.66 | 7.75 | 145.45 |
| 2012 | 63.28 | 17.56 | 62.02 | 19.81 | 0.19 | 182.54 |
| 2013 | 71.60 | 19.99 | 69.04 | 23.01 | 0.00 | 186.98 |
| 2014 | 71.28 | 15.92 | 69.64 | 19.41 | 6.91 | 150.06 |
| 2015 | 71.69 | 17.33 | 69.33 | 21.35 | 2.09 | 177.66 |
| 2016 | 64.46 | 17.90 | 60.38 | 20.44 | 0.86 | 162.83 |
| 2017 | 63.17 | 17.04 | 60.39 | 18.88 | 12.31 | 246.06 |
| 2018 | 71.78 | 22.37 | 67.24 | 32.10 | 16.28 | 268.69 |

Table S5: Mean, standard deviation (sd), median, interquartile range (IQR), min and max of daily median of relative humidity [%] exposure across England during summers 2007-2018.

| year | mean | sd | median | IQR | min | max |
|---|---|---|---|---|---|---|
| 2007 | 0.94 | 0.05 | 0.95 | 0.08 | 0.49 | 1.00 |
| 2008 | 0.93 | 0.06 | 0.95 | 0.09 | 0.51 | 1.00 |
| 2009 | 0.93 | 0.06 | 0.94 | 0.09 | 0.43 | 1.00 |
| 2010 | 0.91 | 0.07 | 0.93 | 0.12 | 0.46 | 1.00 |
| 2011 | 0.92 | 0.06 | 0.93 | 0.09 | 0.50 | 1.00 |
| 2012 | 0.96 | 0.05 | 0.97 | 0.06 | 0.54 | 1.00 |
| 2013 | 0.91 | 0.07 | 0.92 | 0.10 | 0.45 | 1.00 |
| 2014 | 0.91 | 0.06 | 0.91 | 0.09 | 0.50 | 1.00 |
| 2015 | 0.90 | 0.07 | 0.91 | 0.11 | 0.47 | 1.00 |
| 2016 | 0.94 | 0.06 | 0.95 | 0.08 | 0.50 | 1.00 |
| 2017 | 0.93 | 0.06 | 0.94 | 0.08 | 0.49 | 1.00 |
| 2018 | 0.88 | 0.09 | 0.90 | 0.14 | 0.37 | 1.00 |

Table S6: Percentage hospitalisation risk of COPD for every 1$^o$C increase in summer temperature using the model adjusted for relative humidity and national holidays and the different temperature thresholds $c$.

| quantile | threshold $c$ ($^o$C) | WAIC | Effect bellow $c$ | Effect above $c$ |
|---|---|---|---|---|
| 50 | 20.6 | 2,257,389 | 0.12 (-0.22, 0.46) | 1.56 (1.26, 1.84) |
| 55 | 21.0 | 2,257,373 | 0.12 (-0.23, 0.48) | 1.56 (1.26, 1.86) |
| 60 | 21.3 | 2,257,371 | 0.16 (-0.21, 0.49) | 1.53 (1.24, 1.81) |
| 65 | 21.7 | 2,257,379 | 0.21 (-0.12, 0.57) | 1.50 (1.21, 1.78) |
| 70 | 22.1 | 2,257,333 | 0.22 (0.10, 0.55) | 1.49 (1.22, 1.77) |
| 75 | 22.6 | 2,257,331 | 0.37 (0.05, 0.65) | 1.42 (1.15, 1.68) |
| 80 | 23.2 | 2,257,273 | 0.37 (0.09, 0.65) | 1.46 (1.19, 1.71) |
| 85 | 23.8 | 2,257,330 | 0.44 (0.18, 0.70) | 1.46 (1.20, 1.72) |
| 90 | 24.9 | 2,257,440 | 0.62 (0.38, 0.86) | 1.39 (1.11, 1.66) |
| 95 | 26.5 | 2,257,351 | 0.72 (0.49, 0.93) | 1.50 (1.20, 1.82) |

Table S7: Median and 95% credible intervals of the % risk COPD hospitalisation for every $1^o$ increase in warm temperatures by age group and sex. RH+NL refers to adjustment for relative humidity and national holidays, whereas RH+NL+POL for additional adjustment for $PM_{2.5}$ and $O_3$.

| Sex | Age group | Unadjusted models | RH+NL | RH+NL+POL |
|---|---|---|---|---|
| Males | 0-64 | 0.97 (0.24, 1.73) | 1.32 (0.57, 2.06) | 0.64 (-0.24, 1.49) |
| Females | 0-64 | 0.92 (0.25, 1.63) | 1.14 (0.39, 1.84) | -0.04 (-0.90, 0.84) |
| Total | 0-64 | 1.00 (0.51, 1.48) | 1.28 (0.75, 1.82) | 0.35 (-0.26, 0.98) |
| Males | 65-74 | 1.13 (0.50, 1.71) | 1.39 (0.74, 2.03) | 0.62 (-0.15, 1.39) |
| Females | 65-74 | 1.56 (0.94, 2.20) | 1.75 (1.13, 2.41) | 0.76 ( 0.01, 1.52) |
| Total | 65-74 | 1.31 (0.83, 1.70) | 1.54 (1.07, 2.03) | 0.66 ( 0.13, 1.22) |
| Males | >75 | 1.41 (0.87, 1.91) | 1.51 (0.98, 2.07) | 0.25 (-0.35, 0.92) |
| Females | >75 | 1.29 (0.79, 1.83) | 1.51 (0.94, 2.06) | 0.47 (-0.14, 1.11) |
| Total | >75 | 1.36 (0.96, 1.71) | 1.49 (1.11, 1.89) | 0.38 (-0.10, 0.84) |
| Males | Total | 1.23 (0.89, 1.59) | 1.45 (1.10, 1.83) | 0.51 ( 0.07, 0.94) |
| Females | Total | 1.26 (0.93, 1.62) | 1.46 (1.09, 1.82) | 0.41 (-0.03, 0.84) |
| Total | Total | 1.24 (0.98, 1.51) | 1.47 (1.19, 1.73) | 0.47 ( 0.16, 0.75) |

Figure S1: Boundaries of the 326 Lower Tier Local Authorities of England in 2015 (median size: 208km$^2$).

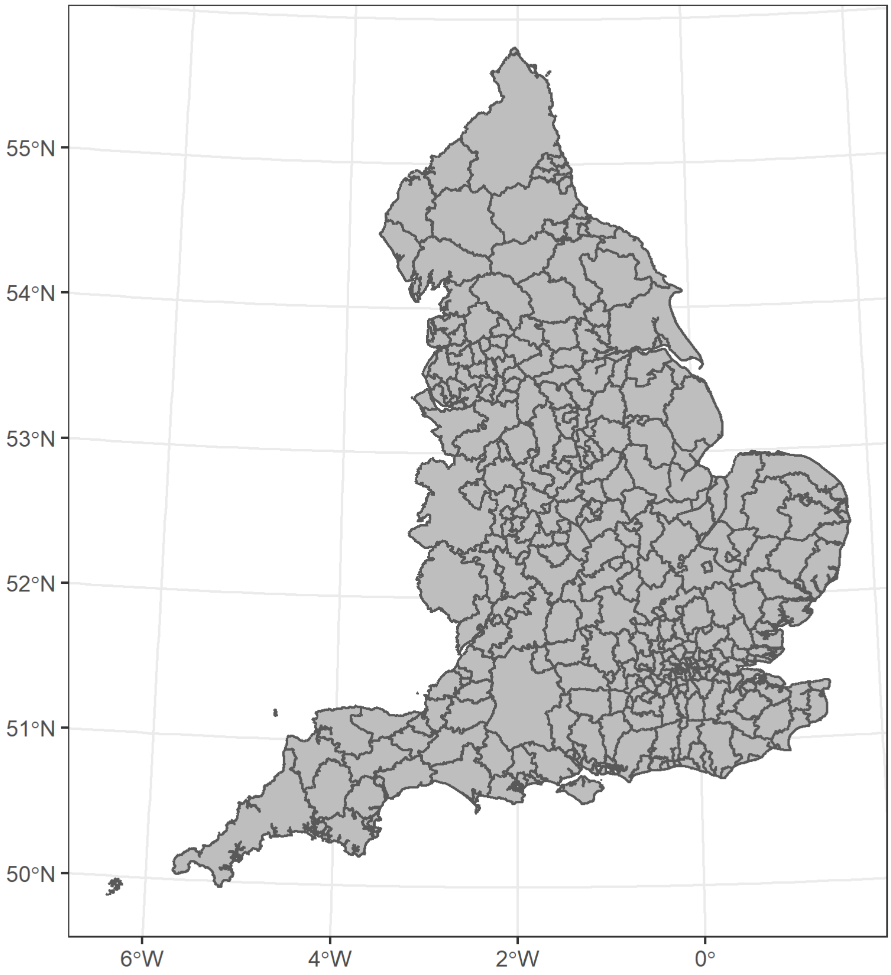

Figure S2: The spatial distribution of the index of multiple deprivation using quintiles in 2015 in England at the lower tier local authority level. Q1 indicates the most deprived areas.

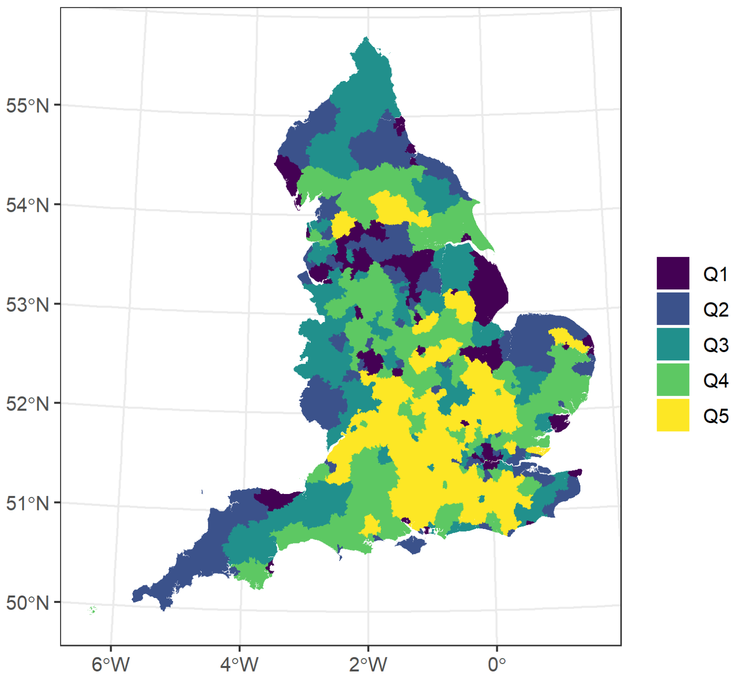

Figure S3: The spatial distribution of urbanicity in based on the Office for National Statistics classification in 2011 at the lower tier local authority level.

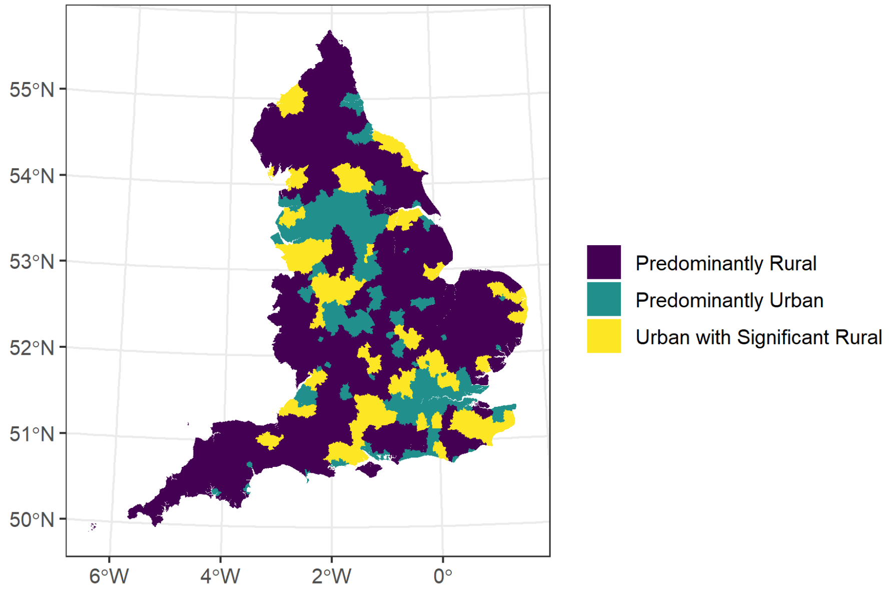

Figure S4: The quintiles of the spatial distribution of the proportion of a lower tier local authority that is covered by green land such as woodland, agricultural land, grassland and other natural vegetated land as classified in the Land Cover Map 2015.

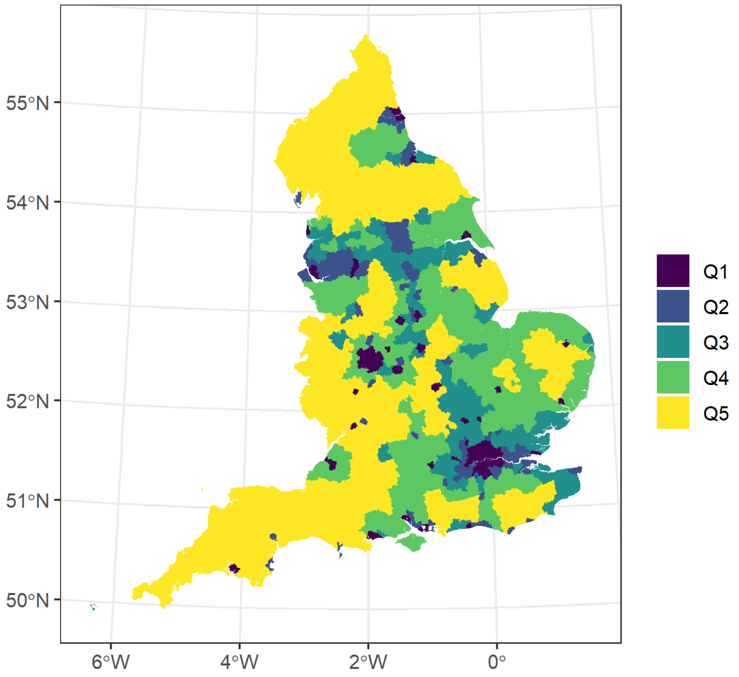

Figure S5: The spatial distribution of the average temperature [$^oC$] by lower tier local authority during 2007-2018 in England.

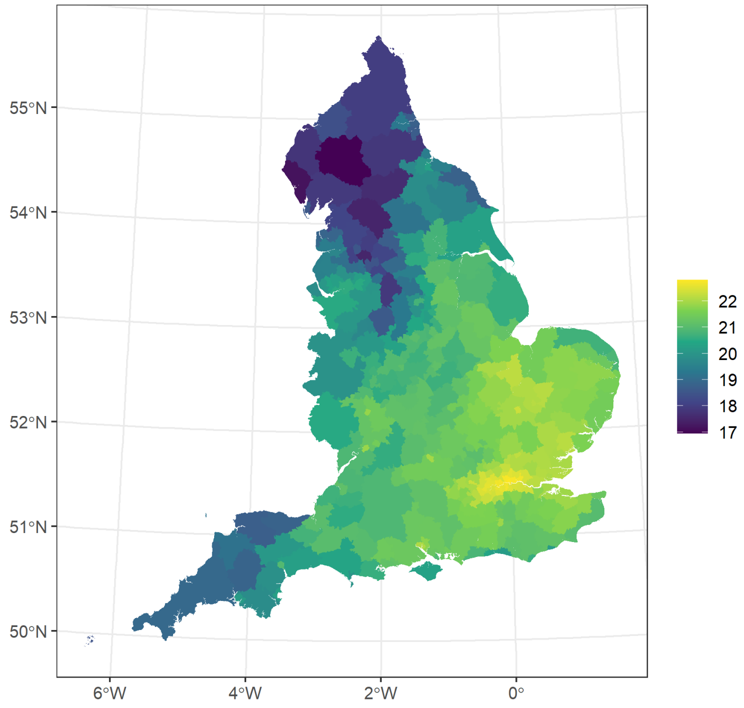

Figure S6: Relative hospitalisation risk (relative to the risk at $18^oC$) using 3rd degree of b-splines at 3 knots and the model with total age and sex and adjusted for national holidays and relative humidity. The red dashed line indicated the threshold $c$ used throughout the study.

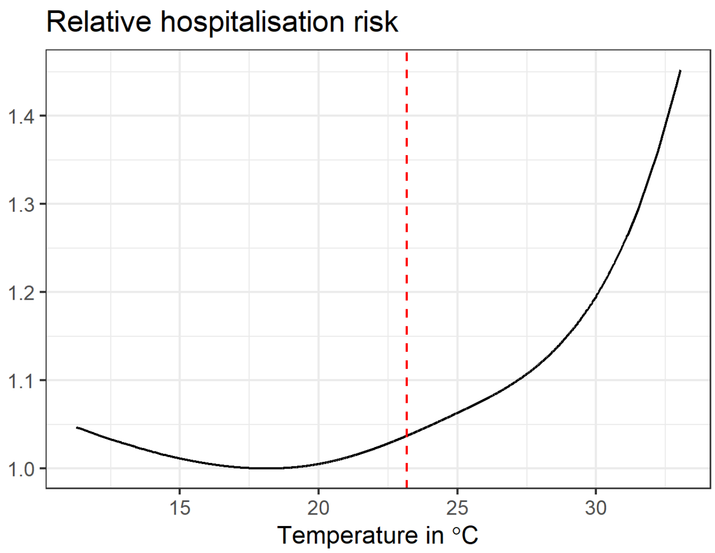
